# A Study on Survival Scenario of COVID-19 patients in India: An Application of Survival Analysis on patient demographics

**DOI:** 10.1101/2020.08.01.20162115

**Authors:** Sampurna Kundu, Kirti, Debarghya Mandal

## Abstract

The study of transmission dynamics of COVID-19, have depicted the rate, patterns and predictions of the pandemic cases. In order to combat the disease transmission in India, the Government had declared lockdown on the 25^th^ of March. Even after a strict lockdown nationwide, the cases are increasing and have crossed 4.5 lakh positive cases. A positive point to be noted amongst all that the recovered cases are slowly exceeding the active cases. The survival of the patients, taking death as the event that varies over age groups and gender wise is noteworthy. This study aims in carrying out a survival analysis to establish the variability in survivorship among age groups and sex, at different levels, that is, national, state and district level. The open database of COVID-19 tracker (covid19india.org) of India has been utilized to fulfill the objectives of the study. The study period has been taken from the beginning of the first case which was on 30^th^ Jan 2020 till 30^th^ June. Due to the amount of under-reporting of data and dropping missing columns a total of 26,815 sample patients were considered. The entry point of each patient is different and event of interest is death in the study. Kaplan Meier survival estimation, Cox proportional hazard model and multilevel survival model has been used to perform survival analysis. Kaplan Meier survival function, shows that the probability of survival has been declining during the study period of five months. A significant variability has been observed in the age groups, as evident from all the survival estimates, with increasing age the risk of dying from COVID-19 increases. When Western and Central India show ever decreasing survival rate in the framed time period then Eastern, North Eastern and Southern India shows a slightly better picture in terms of survival. Maharashtra, Gujarat, Delhi, Rajasthan and West bengal showed alrmingly poor survival as well. This study has depicted a grave scenario of gradation of ever decreasing survival rates in various regions and shows the variability by age and gender.

## 1. Introduction

The whole world is literally perplexed at the sudden outbreak of covid 19 because human race has no remedial measures to combat its lethal impacts. According to the World Health Organisation (WHO), global pandemic covid-19 is derived from a large family of viruses, named the coronavirus that causes respiratory infections which ranges from common cold to high fever, leading to diseases. This blue planet witnessed many epidemic like the acute respiratory syndrome coronavirus during 2002-2003 and H1N1 influenza during 2009 (Cascella et al, 2020) due to various pernicious virus in last two decades but COVID-19 is absolutely incomparable with its ancestors because of its indomitable growth rate and its fatality as well. Though China which was the first to hit the most cases in the beginning, has presently flattened the curve by continuous testing and aggressive quarantine measures. Outside China, South Korea being the country which had the largest initial outbreak has managed to slow down the spread and flatten the curve without imposing lockdown in the country. Their only way of slowing and containing outbreak was mass diagnostic testing and quarantining. It was declared by WHO, to incorporate self-isolation, sanitizing, washing of hands repeatedly and abstaining from touching mouth, face or nose (WHO, 2020). In order to combat the disease transmission in India, the Government had declared lockdown on the 25^th^ of March. Yet the disease has spread like fire all over the country, and as of 30^th^ June, there are 1385494 Cases with 32096 deaths and 886235 recoveries (covid19india.org). For a developing country like India, the pandemic is seriously a big pitfall for the nation, the main sufferers being the marginalized sections of the society. Even after a strict lockdown nationwide, the cases are increasing and have crossed 4.5 lakhs positive cases. Though the fatality rates are less and several studies have shown that the lockdown did slowdown the rate of increase in the cases (Dwivedi et al., 2020). A positive point to be noted amongst all that he recovered cases are slowly exceeding the active cases.

In the study of dynamics of infectious diseases, the compartmental models and basic reproductive number have been seen to be mostly used over the year (Delamater et al., 2019). The basic mathematical models like the Gompertz, Exponential or Logistic growth models have shown to be quite effective in understanding the patterns of the growth of the disease (Ram & Kumar, 2020). One of the main demerits of the Indian database for covid-19 is the under-reporting of the cases due to misreporting and also less number of testing (Dhillon et al., 2020). Amidst the growing number of deaths, researchers all around the world have associated additional important co-factors which are effect on elderly population and impact of pollution, smoking as well as development of acute respiratory distress syndrome (Conticini et al, 2020, Xu et al, 2020). The district level analysis from a study have shown that 92 districts are in red zones of the disease (Unisa et al., 2020). Mostly these red zones are found in the states Maharashtra nd Gujarat, as predicted by the ARIMA model also in study that the cases will be increasing alarmingly (Tyagi et al., 2020).

Studies have shown the impact of lockdown, transmission dynamics of the disease and forecasts. The survival of the patients, taking death as the event that varies over age groups and gender wise is noteworthy. This study aims in carrying out a survival analysis to establish the variability in survivorship among age groups and sex, at different levels, that is, national, state, district and patient level. This is an exceptional quantitative analysis (with analysis of the patient data of COVID-19) not only for its gravity or pertinence but also for its subtle nuances and penetrating approach.

## 2. Data and Methods

Data for the present study has been retrieved from the data sharing portal of India, that is, covid19india.org. The patient level data has been used for the study, which consist of the time to event data. Here the study period is from the beginning of the first case which was on 30^th^ Jan 2020 till 30^th^ June that i.e., five months or 150 days. The entry point of each patient is different and event of interest is death in the study. If the event has not occurred then, the survival time is taken to be censored. Due to the amount of under-reporting of data and dropping missing columns a total of 26,815 sample patients were considered. The inclusion criteria for each patient being, the date of testing positive, date of change of status, reported age and gender. Survival time has been computed by taking the difference between the date of testing positive for the infection and the date of change of status for each sample patients.

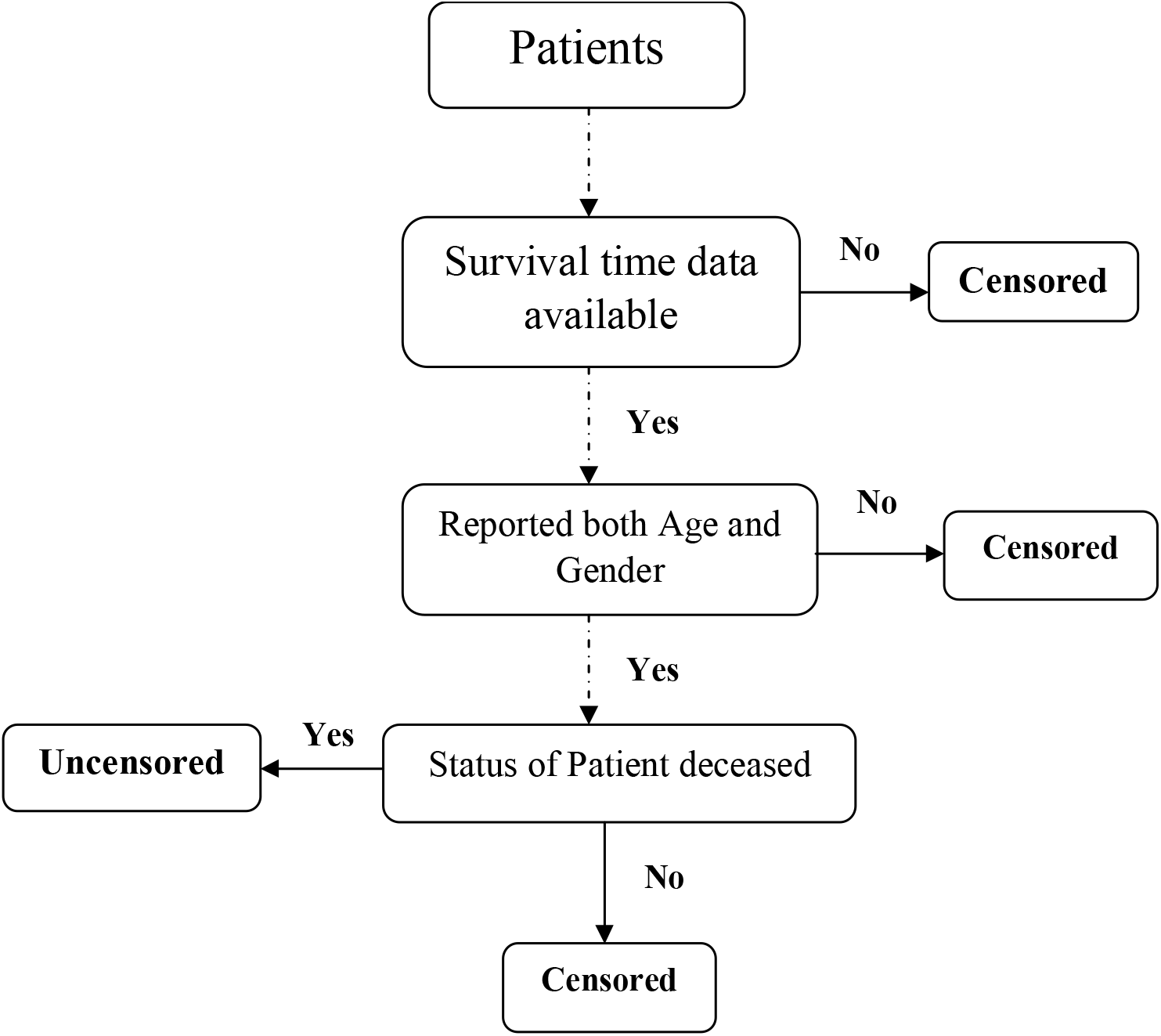

Kaplan Meier Survival Estimator method, Cox Proportional Hazard model and multilevel survival model has been used to perform survival analysis. Firstly, Kaplan Meier Survival Estimator method has been used to estimate the survival function from the survival data. To compare the survival functions for different groups i.e. by gender, by different age groups and by various regions, Log-rank test and Gehan-Breslow-Wilcoxon test has been used. For estimating the survival function in presence of various covariates Cox-Proportionate Hazard model has been used with gender, age and regions being the covariates assuming that the hazard is independent of time. Cox-proportionate hazard model will be as follows:

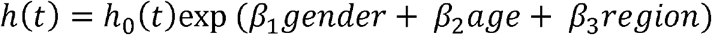

where,

*t* represents the survival time, *h(t)* is the hazard function determined by a set of 3 covariates (gender, age, region). The coefficients (*β*_*1*_, *β*_*2*_, *β*_*3*_) measure the impact (i.e., the effect size) of covariates. The term *h(0)* is called the baseline hazard, that corresponds to the value of the hazard if all the xi’s are equal to zero.

Lastly, Multilevel mixed effects survival analysis has been done as clustering of lower level units at higher level units is a common scenario in such studies. In here it can be clustering of patients at district level, then districts are clustered at state level and all the states at national level. We consider *i*-1,2,…..,*N* clusters (e.g, states, districts) with each cluster having *j=1,2,…,n*_*i*_ patients. Let *S*_*ij*_ be the true survival time of the *j*^*th*^ patient in the *i*^*th*^ cluster, *T*_*ij*_=min(*S*_*ij*_,*C*_*ij*_) be the observed survival time, and *C*_*ij*_ be the censoring time. The proportional-hazards mixed-effects survival model can be written as

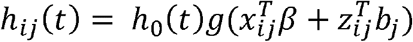

where *h*_*0*_*(t)* is the baseline hazard function of a standard parametric model, such as here we use Weibull at each level because, according to the Akaike’s Information Criterion it is the most appropriate model to be used in comparison to the other models. Therefore, a three level cluster analysis will help in eliminating the variability at each level due to inter correlation between the units and can obtain better estimates of the survival function.

## 3. Results

The Kaplan Meier estimates have been obtained in the beginning to estimate the survival functions by gender, age groups and region. We can see from **Figure 1**, that the survival curves from KM estimator for male and female are almost the same. In **Table 1**, it can be observed that Log rank test (*p*=0.203) and Wilcoxon test (*p*=0.107) for comparing the survival function, both are not significant indicating that there is no significant difference between the survival curves of male and female.

**Figure 1.**
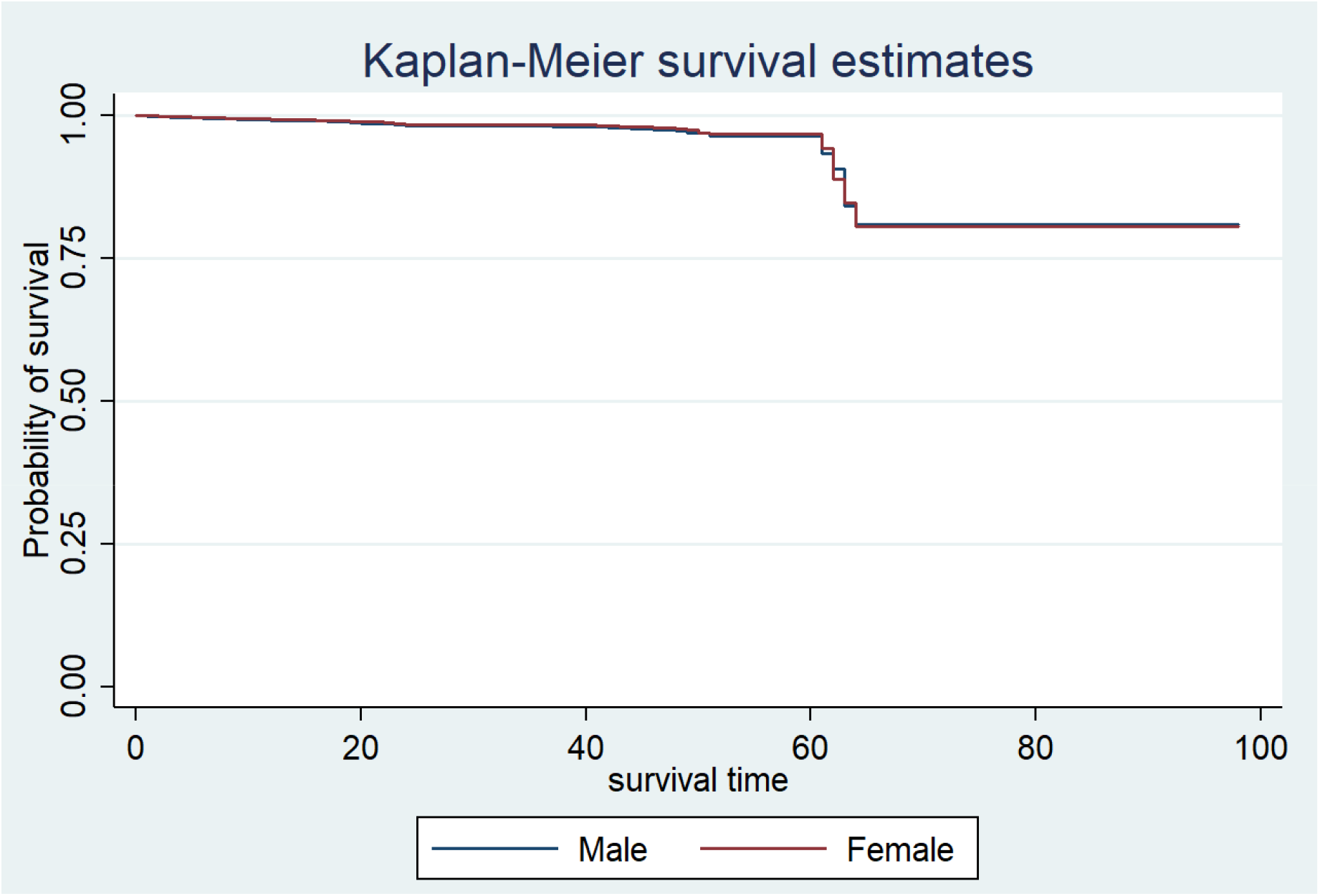
Kaplan Meier Estimate by Gender.

**Table 1:** Kaplan-Meier Estimator: Comparison of Survival Functions by Gender.

|  |  |  |  |
| --- | --- | --- | --- |
| *Test* | *Chi-Square* | *Degrees of Freedom* | *p-value* |
| Log Rank | 1.62 | 1 | 0.203 |
| Wilcox | 2.61 | 1 | 0.107 |

As seen from **Figure 2**, that the survival curves from Kaplan Meier estimator by five year age groups are significantly different. This result is further supplemented by Log rank test (*p*<0.001)and Wilcoxon test (*p*<0.001) for comparing the survival functions, in **Table 2**, which shows that both tests are highly significant indicating that there are significant differences among the survival curves by various age groups.

**Figure 2.**
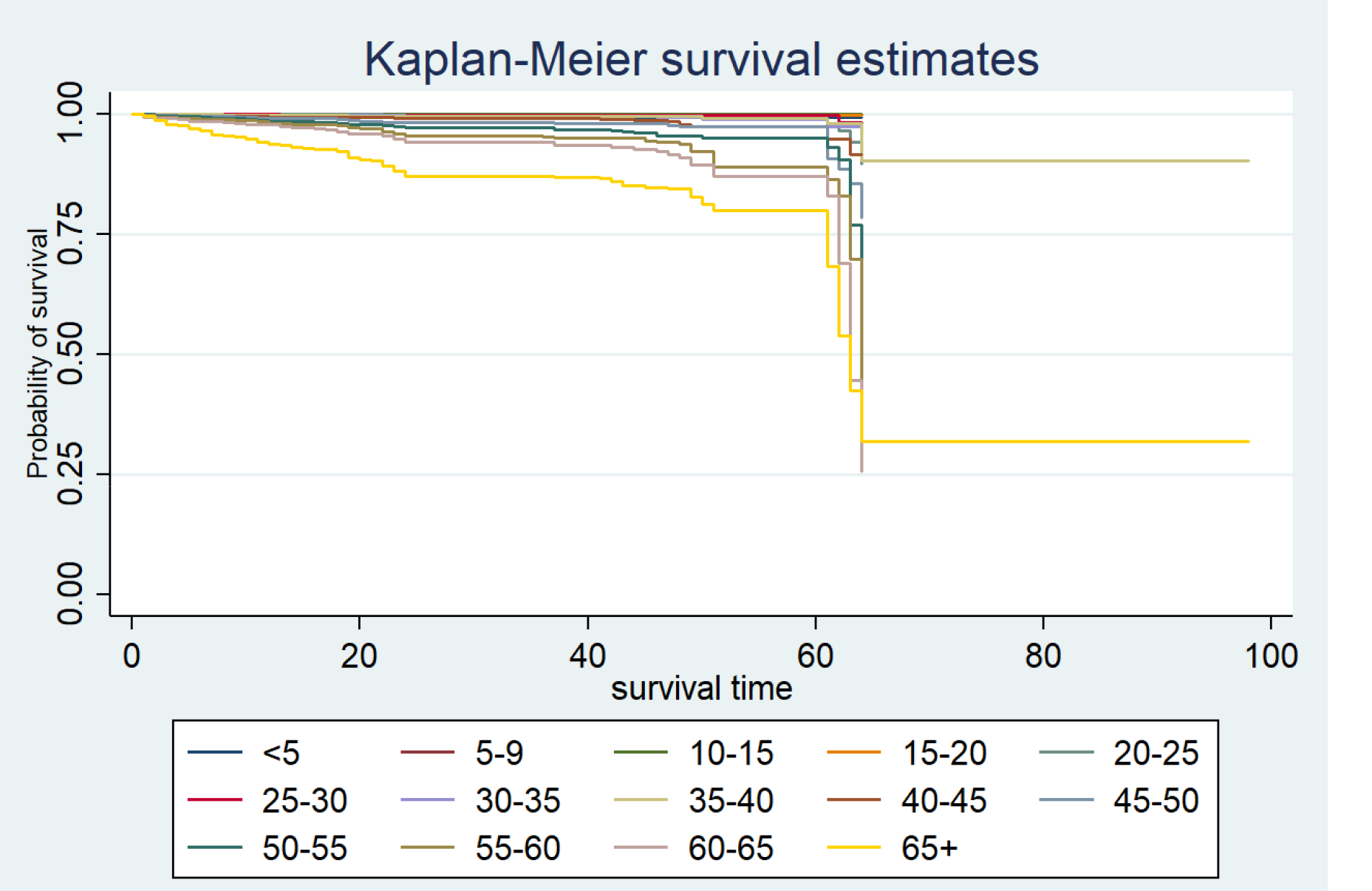
Kaplan Meier Estimate by Age group.

**Table 2:** Kaplan-Meier Estimator: Comparison of Survival Functions by Age Groups.

|  |  |  |  |
| --- | --- | --- | --- |
| *Test* | *Chi-Square* | *Degrees of Freedom* | *p-value* |
| Log Rank | 1302.79 | 13 | <0.001 |
| Wilcox | 1072.26 | 13 | <0.001 |

**Figure 3** depicts that the survival curves from Kaplan Meier estimator by different regions of India are significantly different. From **Table 3**, we infer that Log rank test (*p*<0.001) and Wilcoxon test (*p*<0.001) for comparing the survival functions, both are highly significant indicating that there are significant differences among the survival curves by regions due to regional variations. Therefore, we find that both age and region are significantly associated with the survival rate of COVID-19, without adjusting for other covariates. **Figure 4** depicts the comparison of survival curves among the most affected states by COVID-19, which shows the poor survival in Maharashtra, Delhi, Gujarat, West Bengal and Rajasthan.

**Figure 3.**
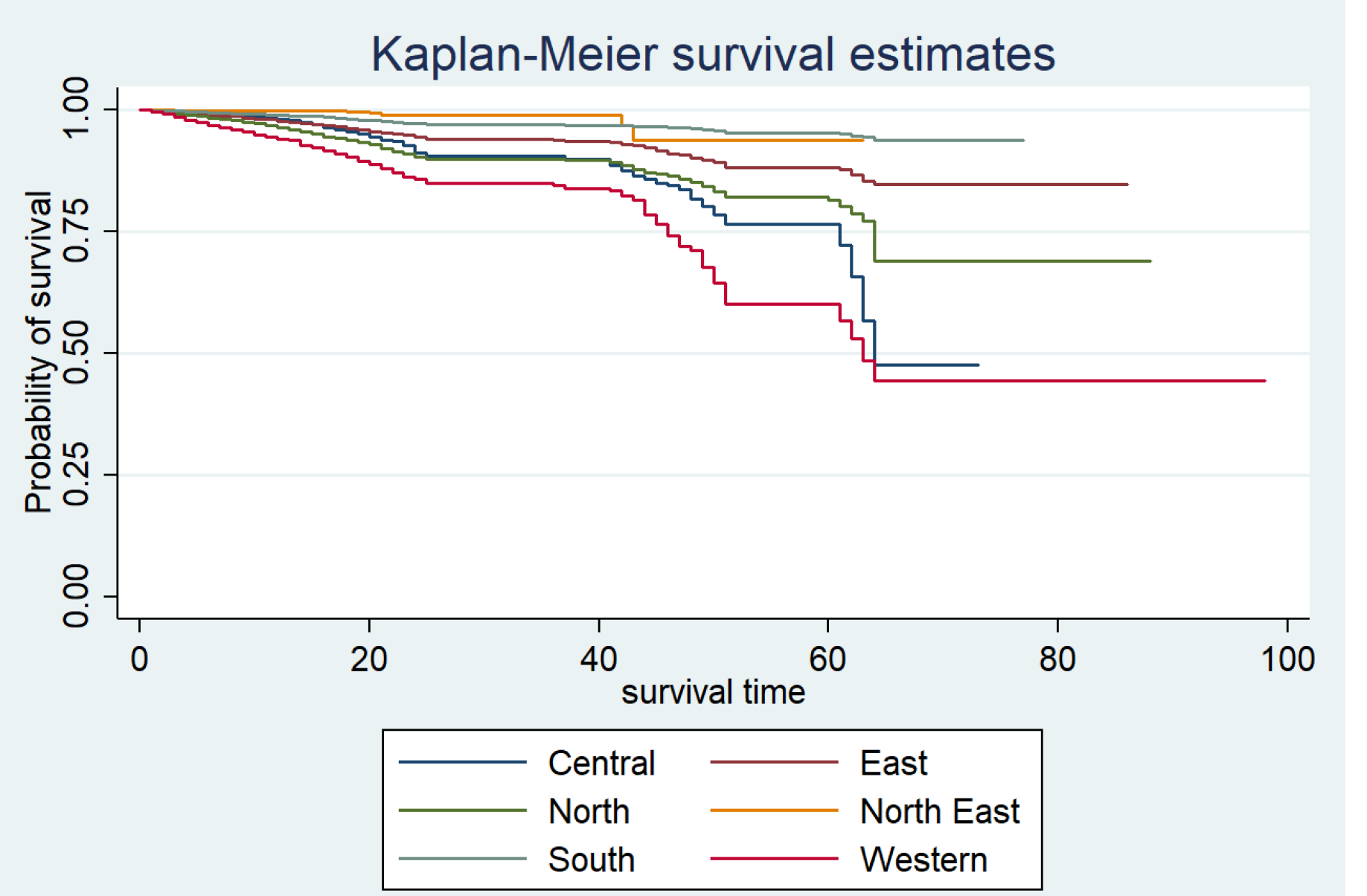
Kaplan-Meier survival estimated by region.

**Figure 4.**
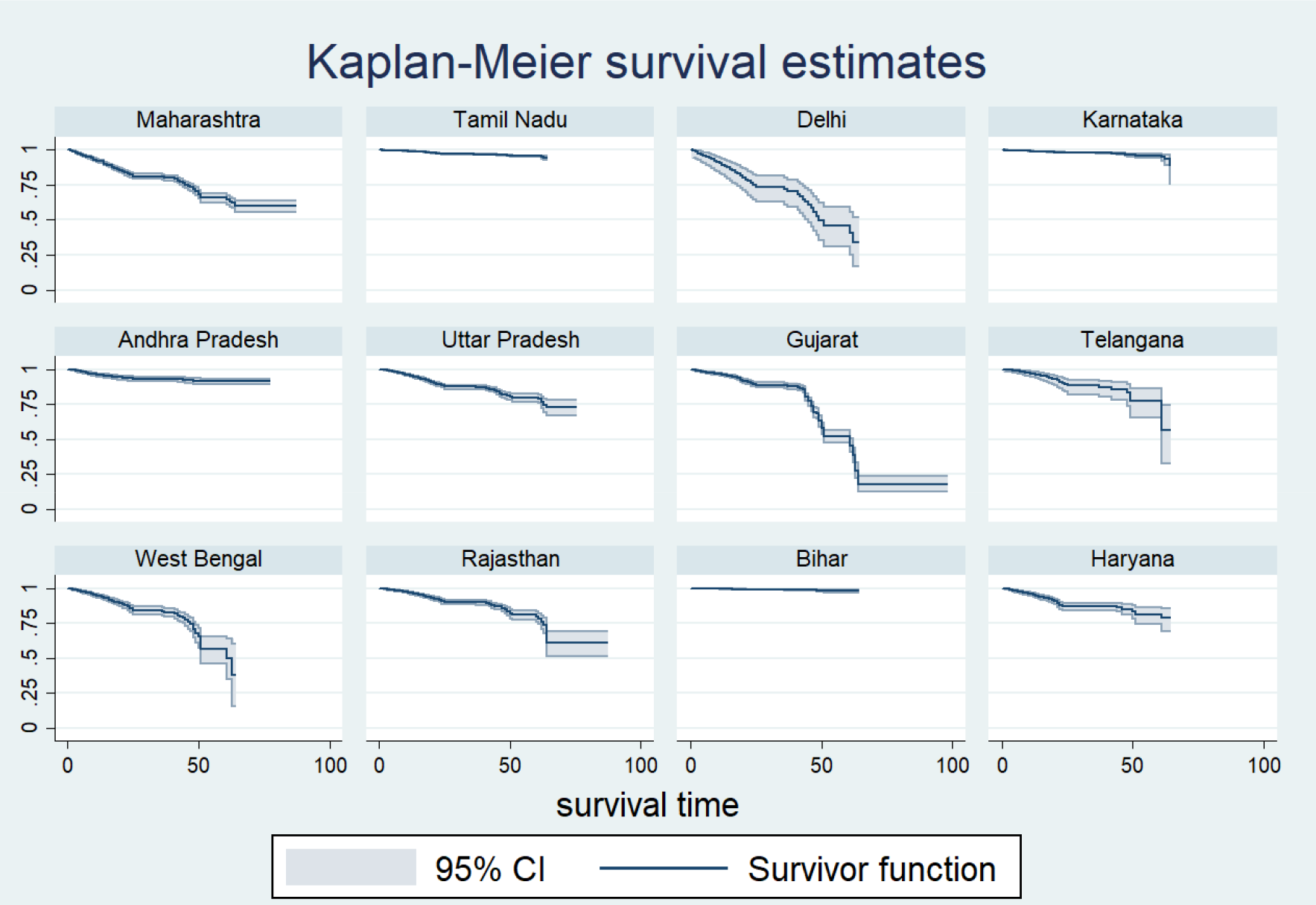
Kaplan-Meier survival Estimates for Most Affected States.

**Table 3:** Kaplan-Meier Estimator: Comparison of Survival Functions by region.

| **Table 3: Kaplan-Meier Estimator: Comparison of Survival Functions by Regions** |  |  |  |
| --- | --- | --- | --- |
| *Test* | *Chi-Square* | *Degrees of Freedom* | *p-value* |
| Log Rank | 1997.43 | 5 | <0.001 |
| Wilcox | 1175.27 | 5 | <0.001 |

**Table 4** represents results of the survival analysis using Cox Proportional Hazard model reiterates that males patients of COVID-19 have 1.14 times more risk of dying as compared to the female patients (HR:1.14; SE:0.11; CI: 0.93,1.38). Coming to the age-wise comparison, we observe that from 45-49 till 65 and above, are having 5.83, 10.08, 15.31, 22.03, 39.21 times higher risk of dying due to COVID-19, as compared to those in the age-group 0-5 years, respectively. The highest risk of death from the disease was among people in the 65+ age group, with 39.2 times higher risk (*p*<0.001), but with a larger confidence interval (CI: 9.73,157.97). While, analyzing survival curve by regions, we can see that East, North East and South Indian regions are at 59%, 14% and 26% less risk of dying from COVID-19 infection as compared to Central India, whereas West Indian region is having 1.9 times more risk of dying as compared to Central India.

**Table 4:**
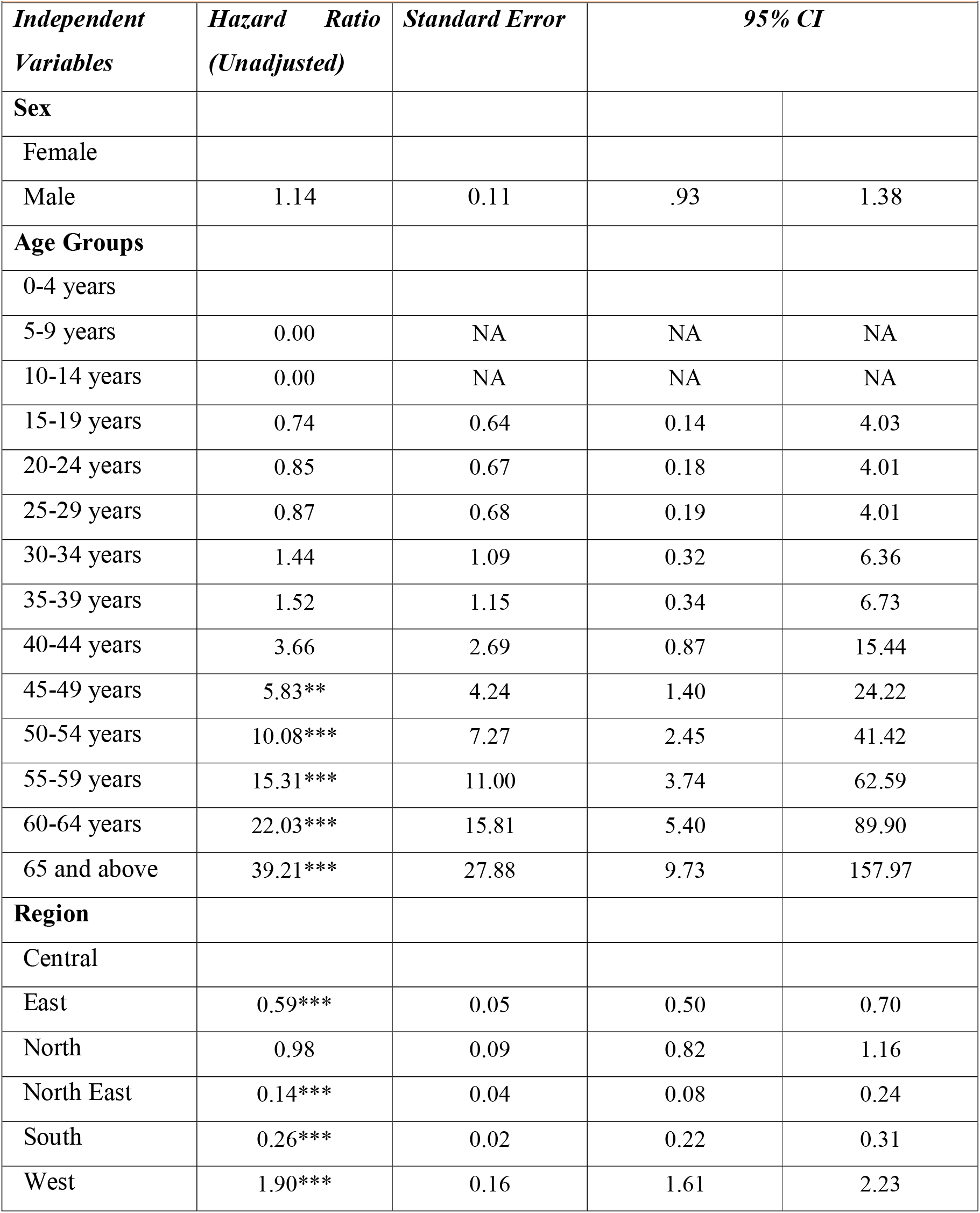
Cox Proportionate Hazard Model Showing UnadjustedHazard Ratios and 95% Confidence Intervals for the deaths occurring due to COVI-19 in India.

| *Independent Variables* | *Hazard Ratio (Unadjusted)* | *Standard Error* | *95% CI* |  |
| --- | --- | --- | --- | --- |
| **Sex** |  |  |  |  |
| Female |  |  |  |  |
| Male | 1.14 | 0.11 | .93 | 1.38 |
| **Age Groups** |  |  |  |  |
| 0-4 years |  |  |  |  |
| 5-9 years | 0.00 | NA | NA | NA |
| 10-14 years | 0.00 | NA | NA | NA |
| 15-19 years | 0.74 | 0.64 | 0.14 | 4.03 |
| 20-24 years | 0.85 | 0.67 | 0.18 | 4.01 |
| 25-29 years | 0.87 | 0.68 | 0.19 | 4.01 |
| 30-34 years | 1.44 | 1.09 | 0.32 | 6.36 |
| 35-39 years | 1.52 | 1.15 | 0.34 | 6.73 |
| 40-44 years | 3.66 | 2.69 | 0.87 | 15.44 |
| 45-49 years | 5.83** | 4.24 | 1.40 | 24.22 |
| 50-54 years | 10.08*** | 7.27 | 2.45 | 41.42 |
| 55-59 years | 15.31*** | 11.00 | 3.74 | 62.59 |
| 60-64 years | 22.03*** | 15.81 | 5.40 | 89.90 |
| 65 and above | 39.21*** | 27.88 | 9.73 | 157.97 |
| **Region** |  |  |  |  |
| Central |  |  |  |  |
| East | 0.59*** | 0.05 | 0.50 | 0.70 |
| North | 0.98 | 0.09 | 0.82 | 1.16 |
| North East | 0.14*** | 0.04 | 0.08 | 0.24 |
| South | 0.26*** | 0.02 | 0.22 | 0.31 |
| West | 1.90*** | 0.16 | 1.61 | 2.23 |

A multi-level survival model has been carried out at overall national level (Weibull Regression), state and district level (Mixed effects Weibull Regression), taking age-groups and gender as the covariates. The results of the multilevel survival analysis, that is after controlling variability due to clustering of lower level at the higher level, it can be seen from **Table 5**, that hazard ratio at India level shows that, males are 1.27 times at higher risk that female patients of dying from COVID-19 (HR:1.27; SE:0.13), which is almost the same at state (HR:1.32; SE:0.13) as well as district (HR:1.21; SE:0.13) level. The significant variability in survival has been observed at the age groups more than 45 years. At all levels, we find that the hazard ratio increases with the increasing age groups, but decreases across each level. For instance, in 65+ age group, overall the patient is at 39.3 times higher risk of dying than a patient at younger age group (HR: 39.3; SE:27.94); whereas in state level the hazard ratio is 32.28 and district level is 23.55. Now, from the variance of the errors of model, we can infer that the heterogeneity is more at the district level (σ_e_ ^2^: 6.85; SE: 1.35) than the state level (σ_e_^2^: 2.28; SE: 0.83).

**Table 5:** Multilevel Survival Analysis: By National, State and District level Hazard Ratio for deaths among COVID-19 patients in India.

| *Independent Variables* | *National Level Hazard Ratio(S.E)* | *State Level Hazard Ratio(S.E)* | *District Level Hazard Ratio(S.E)* |
| --- | --- | --- | --- |
| **Sex** |  |  |  |
| Female |  |  |  |
| Male | 1.27 (0.13)** | 1.32 (0.13)** | 1.21 (0.13) |
| **Age Groups** |  |  |  |
| 0-4 years |  |  |  |
| 5-9 years | 0.00(0.00) | 0.00 (0.00) | 0.00 (0.00) |
| 10-14 years | 0.00(0.00) | 0.00 (0.00) | 0.00 (0.00) |
| 15-19 years | 0.71 (0.62) | 0.86 (0.75) | 0.49 (0.45) |
| 20-24 years | 0.80 (0.63) | 0.84 (0.66) | 0.68 (0.54) |
| 25-29 years | 0.81 (0.63) | 0.91 (0.71) | 0.71 (0.56) |
| 30-34 years | 1.26 (0.96) | 1.53 (1.16) | 1.22 (0.93) |
| 35-39 years | 1.22 (0.94) | 1.37 (1.05) | 1.13 (0.87) |
| 40-44 years | 3.36 (2.47) | 4.00 (2.94) | 3.37 (2.49) |
| 45-49 years | 5.20 (3.78)** | 6.22 (4.52)** | 4.89 (3.56)** |
| 50-54 years | 8.53 (6.16)*** | 10.70 (7.73)*** | 7.77 (5.63)** |
| 55-59 years | 13.99 (10.06)*** | 14.76 (10.62)*** | 11.49 (8.29)*** |
| 60-64 years | 20.52 (14.74)*** | 18.40 (13.23)*** | 14.19 (10.24)*** |
| 65 and above | 39.30 (27.94)*** | 32.28 (23.00)*** | 23.55 (16.84)*** |
| **Error Variance** |  | 2.28(0.83) | 6.85(1.35) |

## 4. Discussion

This study strived to find out reliable features associated with survival patterns and inevitably scanned the role of gender, age and regional variability as controlling factors of survival rate. For the survival analysis, study period was of five months with death being the event of our analysis. While evaluating the Kaplan Meier survival function, it has been observed that the probability of survival has been declining during the study period of five months. In the midst of the study period no stabilization can be observed. Female were found to have better survival as compared to their counterparts, evident from the Cox PH results, which might be due to sex-differentials in cellular compositions and immunological micro-environment of lungs (Wei et al, 2020). Even though there is no such difference in survival curves of male and female, except for a miniscule of difference; it has been stated in earlier studies as well that men with COVID-19 are at higher risk of death and health outcomes, independent of age (Jin et al., 2020) as men are having more burden of diseases (diabetes, hypertension or cardiovascular diseases) therefore, men have shown markedly increased risk of developing severe cases in comparison to women. Also amajor proportion of the confirmed cases is males than females which is expected in the country with the gender hegemony where male work participation, mobility and migration is predominately higher than that of females (Bannerjee & Raju, 2009), which makes men more vulnerable to the infection.

A significant variability has been observed in the age groups, as evident from all the survival estimates, with increasing age the risk of dying from Covid-19 increases (Velavan & Meyer, 2020). It was reported in a study that comorbid COVID-19 patients, nearly 21% had hypertension, 11% had diabetes and 7% had cardiovascular disease(CVD), which increases their risk of mortality (Singh et al., 2020). In contrast to other countries data, in India only 15% of the confirmed cases are above age 60 years and majority of them are from age bracket 25-59, most probably because the older population is the most affected in this pandemic and India has a fairly younger population which might attribute to a lower CFR (Dhillon et al., 2020). Around 84% of the COVID-19 patients were men and 82% patients overall were above 40 years of age, as reported in an ICMR study (Gupta et al, 2020). India is one of the biggest countries of the world and infested with neumerous diversities in every respect. Every province posses its own demographic features typical climatic character and above all own lifestyle. Needless to say these factors play a pivotal role. That’s why variation in survival rate is easily traced. When western and central India show ever decreasing survival rate in the framed time period then eastern, north eastern and southern India shows a slightly better picture in terms of survival. Maharashtra, Gujarat, Delhi, Rajasthan and West bengal showed alrmingly poor survival as well.

## 5. Conclusion

Epitomizing the whole study, it is notable that in the stipulated time period observation has clearly revealed that the survival rate was ever declining and till date that trend is not abated. Worthy to be mentioned that age, gender, regional variability were important determinants at each step. Also, from this study it is crystal clear that due to prevalent co-morbidities and ever nurtured male chauvinism the male population in our country is more vulnerable to COVID-19 (also data supports the fact that survival rate of female population is much higher). This study has also traced a different pattern for India than other countries, as younger population is higher in our country than most of the countries the number of affected people is numerous. Lastly, this study has depicted a grave scenario of gradation of ever decreasing survival rates in various regions. In essence we can safely conclude that critical appraisal of survival rate and thorough analysis of patient data have equipped this study to find out risk group and comparative studies about various segments in India.

## Data Availability

The database is available in the data sharing portal of covid19 India

https://www.covid19india.org/

## Notes

### Competing Interest Statement

The authors have declared no competing interest.

### Funding Statement

No external funding was received in carrying out this study.

